# Cognitive functions supporting learning over time in naming treatment for aphasia

**DOI:** 10.1101/2024.11.25.24317903

**Authors:** Emily B. Goldberg, William D. Hula, Robert Cavanaugh, Alexander M. Swiderski, Alyssa Autenreith, Michael Walsh Dickey

## Abstract

**Purpose:** Aphasia rehabilitation is a learning process that unfolds over time. Previous work has examined aphasia treatment response using pre- to post-treatment comparison, largely ignoring the unfolding learning response that occurs session-to-session. We aimed to: (1) characterize the shape of learning while individuals with aphasia received intensive anomia intervention, and (2) identify the cognitive predictors of this learning response.

**Method:** Individuals (n=39) with chronic post-stroke aphasia received intensive semantic feature analysis (SFA). Naming accuracy for trained and semantically-related, untrained words was probed daily. We used Bayesian generalized linear mixed effects models to estimate the shape of learning during SFA treatment, and to measure the influence of key cognitive functions on treatment response.

**Results:** Most treatment gains appeared early during treatment, after the first four hours of intervention. Verbal recognition and visuospatial memory were associated with the magnitude of those early treatment gains, favoring strong cognitive performers. Treatment generalization to untrained targets was present but modest, with some evidence suggesting that visuospatial recall performance may be associated with treatment generalization.

**Conclusions:** Monitoring SFA treatment response early could help inform clinicians whether patients will respond optimally to intervention. Verbal recognition and visuospatial recall support learning during treatment, helping elucidate cognitive underpinnings of learning during aphasia rehabilitation.

## Introduction

The language and cognitive deficit profiles of individuals with post-stroke aphasia are heterogeneous. However, all individuals with aphasia experience anomia, or word-retrieval difficulty (Laine & Martin, 2006). Because anomia is a pervasive consequence of aphasia and has significant detrimental impact on individuals’ ability to communicate, many speech language pathologists (SLPs) dedicate a substantial amount of treatment time to restoring word-retrieval abilities by implementing established anomia treatment protocols (Tierney-Hendricks et al., 2022). Similarly, many clinical trials aimed at investigating aphasia treatment efficacy have focused specifically on examining efficacies of anomia treatments, such as Semantic Feature Analysis (SFA; Boyle & Coelho, 1995; Massaro & Tompkins, 1992; Quique et al., 2019). However, treatment response in people with aphasia is highly variable, and predicting which patients will respond optimally to SFA or to other widely used aphasia treatments remains a significant challenge faced by the field (Doogan et al., 2018). Given that SFA is the most widely used anomia treatment in clinical speech and language pathology (Raymer & Roitsch, 2023a; Tierney-Hendricks et al., 2022), it is uniquely worthy of detailed study to better understand variability in patient outcomes.

To measure treatment response, majority of group studies have historically presented measures of pre- to post-treatment changes to estimate therapy-driven language improvements (Breitenstein et al., 2017; Edmonds et al., 2014; Kendall et al., 2015; Kristinsson et al., 2021; Stockbridge et al., 2023). In a traditional pre-to-post treatment contrast, performance on a desired outcome measure before treatment is compared to performance on the same measure gathered after treatment. Ideally, at the end of treatment, participants with aphasia will demonstrate a robust and reliable increase or gain in their performance on the desired outcome measure. These pre-post comparisons may involve tests of performance on standardized assessments at a single timepoint before and after treatment in randomized control trials (Brady et al., 2016; Robey, 1998) or they may involve performance on a targeted behavior at multiple timepoints before (at baseline), during, and after treatment (during follow-up) in single-subject experimental designs (Beeson & Robey, 2006).

Using such pre-to-post comparisons, many studies have identified sources of treatment response variability by accounting for contributions of person-specific variables on treatment outcomes. For example, multiple findings indicate that lesion size and site (Hope et al., 2013; Kiran & Thompson, 2019; Plowman et al., 2012; Watila & Balarabe, 2015) and initial aphasia severity (Lazar et al., 2010; Watila & Balarabe, 2015), among other variables, predict the magnitude of change in language function from pre- to post-treatment. Findings from these investigations usefully demonstrate treatment efficacy. However, they do not account for the gradual re-learning process that unfolds during aphasia rehabilitation. Directly examining this relatively less-studied incremental learning process may shed important and novel light on diversity of aphasia treatment responses.

Recently, a handful of investigations among the large aphasia treatment literature have used naming performance gathered ***during*** anomia intervention to consider how within-treatment performance predicts long term patient outcomes. For instance, in a study by Dignam et al. (2023), participants with chronic aphasia engaged in intensive anomia treatment and were probed on their abilities to name trained (i.e. directly treated) and untrained (i.e. never treated) target words after every 3 hours of intervention. The investigators found that performance on the very first naming probe significantly predicted word-retrieval abilities of trained and untrained items, both at the end of treatment and at a follow-up visit that occurred 1 month later. Consistent with these findings, Simic et al. (2020) found that early improvements in anomia treatment, measured via daily naming probes, strongly predicted naming outcomes at the end of treatment. These limited though consistent results suggest that early treatment response predicts anomia treatment outcomes, and that factoring in how treatment response unfolds over time may usefully account for sources of variability in outcomes. Specifically, it appears that rapid treatment responders are more likely to benefit from treatment while slow or absent responders are less likely.

A possible explanation for this advantage is that rapid treatment responders may also be strong learners who are capable of retaining treatment effects in the long-term. Increasingly, aphasia rehabilitation is viewed as a re-learning process (Ferguson, 1999; Hopper & Holland, 2005; Nunn et al., 2023; Raymer et al., 2008) and learning abilities can become impaired in some individuals after stroke, which may predispose them to benefit less from language intervention (Martin & Saffran, 1999; Vallila-Rohter & Kiran, 2013). Learning in the context of aphasia treatment is different from acquisition of novel information, such as learning novel words. Instead of targeting novel information or concepts, aphasia treatment protocols aim to restore access to pre-existing linguistic knowledge through repeated and structured practice. This process requires individuals with aphasia to learn procedural features of the treatment protocol being used, but more importantly also elicits re-learning of their ability to access representations that are inconsistently available for language use (Mirman & Britt, 2014). Although novel learning and the language re-learning are distinct processes, the ability to learn new information after stroke is predictive of aphasia treatment success (Dignam et al., 2016; Tuomiranta et al., 2014). This suggests shared overlapping cognitive substrates (Peñaloza et al., 2022).

Learning ability is a dynamic and complex cognitive skill that relies on synergies of multiple domain-specific non-language cognitive functions, such as memory, attention, and executive functions – cognitive functions that are commonly affected by stroke and have been associated with aphasia treatment outcomes (Diedrichs et al., 2022; Dignam et al., 2017; Gilmore et al., 2019; Lambon Ralph et al., 2010; Seniów et al., 2009). Multiple studies have reported that performance on cognitive tasks – including tasks that measure verbal short-term memory, visuospatial recall, sustained attention, and executive function – predict magnitude of aphasia treatment response (Diedrichs et al., 2022). Specifically, individuals with higher scores on domain-specific cognitive-linguistic measures (indicative of stronger and relatively spared cognitive function) tend to make greater language rehabilitation gains. The cognitive functions that appear to support aphasia treatment success are the same prerequisite cognitive skills required for successful learning of new information, despite these forms of learning representing distinctive processes (Peñaloza et al., 2022).

The exact characterization of re-learning in the context of aphasia treatment is undefined and must be specified more precisely (Nunn et al., 2023). The current study examines this incremental process directly, modeling changes over time during treatment to identify important patterns in the session-to-session variability seen in word re-learning among individuals with chronic post-stroke aphasia.

### Study Aims

The current study had two aims: (i) to characterize the course of word re-learning while individuals with aphasia engage in SFA and (ii) to determine how non-language cognitive function influences the course of re-learning during treatment. The first aim focused on session-by-session learning, and addressed the following research question:

**1.** As people with aphasia actively engage in SFA, what is the shape of the learning curve for trained and untrained target words during treatment?

Based on findings from Dignam et al. (2023) and Simic et al. (2020), we hypothesized that individuals with aphasia would, on average, show rapid improvements in their ability to name trained and untrained target words early in the course of SFA. Additionally, those improvements would be greater for directly trained items than for untrained (Aim 1).

The second aim examined the non-language cognitive predictors of the shape of the learning curve for trained and untrained target words during treatment, addressing the following research question:

**2.** As people with aphasia actively engage in SFA, which non-language cognitive factors are associated with the shape of the learning curve for trained and untrained target words during treatment?

We hypothesized that verbal short term memory, sustained attention, and visuospatial recall abilities would be associated with better re-learning throughout treatment (Diedrichs et al., 2022; Majerus, 2013; Peñaloza et al., 2022; Rodríguez-Fornells et al., 2009; Ullman, 2004; Varkanitsa et al., 2023; Yu & Smith, 2007). Specifically, better treatment responders (individuals either with greater level change over baseline slope or steeper slope change compared to baseline slope) would also be strong performers on these measures of cognitive functions, whereas less robust treatment responders (individuals either with small level change over baseline slope or less steep slope changes compared to baseline slope) would also be weak performers on cognitive measures.

## Methods

The present study is a retrospective analysis of existing data. Methods supporting data collection and analyses received approval from the Veterans Administration Pittsburgh Healthcare System Institutional Review Board. Participants or their legally authorized representatives provided written informed and verbal consent. Study methods are described comprehensively in Evans et al. (2021) and Gravier et al. (2018).

### Participants

Study investigators enrolled 44 individuals with chronic (>6 months post-onset) aphasia following unilateral, left hemisphere stroke. Participants with recurrent stroke and known comorbid neurologic disease other than stroke were excluded. Using the Comprehensive Aphasia Test (CAT; Swinburn et al., 2012), investigators measured language abilities across modalities and aphasia severity to confirm aphasia status and inform study inclusion. Specifically, we generated a CAT modality mean T-score by averaging T scores for the following subtests: spoken and written comprehension, repetition, naming, and reading aloud. Participants with CAT naming modality T-scores of 40 or above were eligible to participate.

While 44 total patients were eligible and completed the study in its entirety, the protocol for the first five research participants deviated from the protocol for the remaining 39 individuals. Specifically, the first five participants were treated on more word lists for longer periods of time. Therefore, we excluded the first five participants to improve internal validity and make analogous comparisons regarding treatment effects, resulting in a final sample of 39 individuals with chronic aphasia (see Evans et al., 2021 for separate analyses of data drawn from the same sample of participants). Table 1 offers a summary of demographic information for the 39-person sample. Means and standard deviations of the sample’s performance on language and non-language cognitive evaluations are also listed in Table 1.

**Table 1.** Summary of demographic information and breakdown of language and non-language cognitive scores for study sample (*N*=39).

| Sample Demographics |  |  |  |
| --- | --- | --- | --- |
| Demographic | Summary Statistic | Range |  |
| Age (years) | 61.8±12.2 | 24-78 |  |
| Education (years) | 14.8±3.04 | 10-25 |  |
| Months Post Onset | 59±56.2 | 6-245 |  |
| Sex | Female: 4 | NA |  |
|  | Male: 35 |  |  |
| Handedness | Left: 3 | NA |  |
|  | Right: 36 |  |  |
| Race | African American: 6 | NA |  |
|  | Hispanic: 1 |  |  |
|  | White: 32 |  |  |
| Sample Cognitive Performance |  |  |  |
| Measure | Mean and SD Total | Max Possible Score | Range |
| CAT Modality Mean T-Score | 52.8±4.74 | NA | 44.3-64.2 |
| Camden Verbal Memory Test:<br>Paired Associate Learning<br>Subtest | 20.8±4.09 | 25 | 10-25 |
| Rey Figure Copy: Immediate<br>Delay | 13.3±6.43 | 36 | 0-30 |

Note. CAT = Comprehensive Aphasia Test.

### Study Timeline

Once determined eligible for inclusion, individuals in our sample engaged in 3 distinct phases of the study: (1) Baseline Testing, (2) Treatment Period, and (3) Post-Treatment Maintenance Period. The **baseline testing** occurred over several days. The active SFA **treatment period** lasted 4 weeks. Immediately following the four-week period of intervention, participants repeated select language measures at the onset of the **post-treatment maintenance period**. Because they are not relevant to the current study, post-treatment maintenance procedures will not be discussed further here. Relevant to the current study are the baseline testing and treatment period phases, which are detailed below.

#### Phase 1: Baseline Testing & Stimulus Development

Immediately prior to intensive SFA treatment, all participants completed an extensive battery that assessed language and non-language cognitive abilities. Language abilities were measured using the CAT and a large 194-item naming battery. All 194 items were named 2 or 3 times during the baseline evaluation phase. Research participants also completed several non-language cognitive measures including the Camden Memory Test (Warrington, 1996), Rey Figure Copy and Immediate Recall (Meyers & Meyers, 1995), and the Test of Everyday Attention (TEA; Robertson et al., 1994). Informed by findings reported in Dignam et al. (2017), Gilmore et al. (2019), Lambon Ralph et al. (2010), Seniów et al. (2009), and Simic et al. (2019), we identified three key non-language cognitive variables of interest, which we used to examine the influence of domain-specific cognitive functions on anomia treatment response: (1) verbal recognition memory, (2) visuospatial recall memory, and (3) selective attention. The standardized test and associated subtest that we used to represent language and non-language cognitive functions are listed in Table 2.

**Table 2.** Measures used to gauge specific language and non-language cognitive functions of interest.

| Cognitive Function | Measure |
| --- | --- |
| Language | • CAT Modality Mean T-Score<br>• 196-item naming battery |
| Verbal Recognition Memory | • Camden Verbal Memory Test: Paired Associate Learning Subtest |
| Visuospatial Recall | • Rey Figure Copy: Immediate Delay |
| Sustained Attention | • TEA: Elevator Count with Distraction |

Treatment stimuli were selected using individual performance on the repeated 194-item naming battery. Specifically, words that a participant named incorrectly on at least 2 of 3 baseline naming battery administrations were identified as potential targets and guided treatment list generation. Three treatment lists were created for every participant, and each list contained 10 words belonging to one of 8 possible semantic categories (animals, birds, fruits & vegetables, musical instruments, occupations, sports equipment, tools, and transportation). If participants incorrectly named multiple nouns across more than 3 semantic categories, their preferences were considered when creating the treatment lists. Each treatment list contained 5 semantically-related words that would be directly targeted during intensive SFA (**trained items**), and 5 semantically-related words that would never be treated during SFA sessions (**untrained items**), which were included to monitor response generalization.

#### Phase 2: Treatment Period

Participants engaged in individually delivered, intensive SFA treatment for four weeks. Treatment sessions occurred twice a day, 4-5 days per week, and lasted 120 minutes each. Thus, it is estimated that participants engaged in approximately 4 treatment hours per day. Licensed SLPs delivered SFA treatment using a computer program designed by Winans-Mitrik and colleagues (2013). For each trial, SLPs showed participants a picture of a trained item and participants attempted to name the item. Participants then generated a minimum of 3 physical properties, actions, and locations, and 1 category and personal association per trial. While participants generated features, the treating clinician recorded responses using a computerized SFA visual organizer (Figure 1), which was visible to the participant.

**Figure 1.**
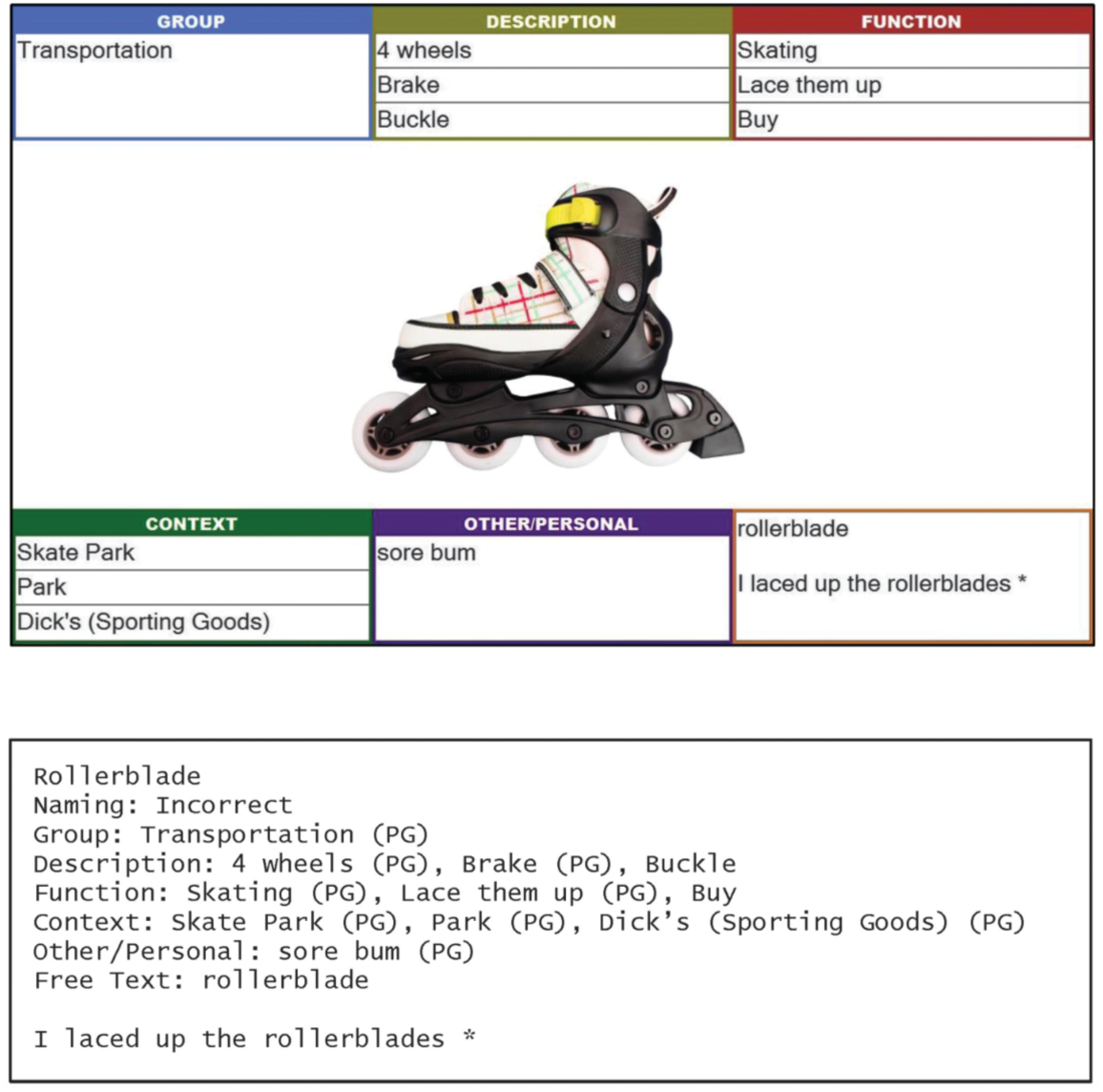
Example of the visual organizer that the clinician and research participant worked through for each SFA trial (Gravier et al., 2018).

If participants had trouble generating features, SLPs implemented a cueing hierarchy. After 10 seconds if no feature was generated, SLPs provided an open-ended and targeted cue (e.g. “what does it feel like?”). If participants continued to struggle with feature generation, the SLP presented binary forced-choice questions (e.g. “is this hard or soft?”). After completing feature generation across category, physical properties, actions, locations, and personal association for a given trained item, the participant attempted to name the target word again. Erroneous or absent responses prompted the clinician to model target word production, eliciting repetition from the participant. Additional details regarding specific decisions made during treatment are outlined in Gravier et al. (2018).

Prior to each treatment session, participants completed a 10-item naming probe. Figure 2 depicts the schedule before, during, and after treatment. These daily naming probes measured naming accuracy of the 5 trained and 5 untrained items comprising the list being actively targeted. When participants achieved 80% naming accuracy or greater on 3 consecutive daily naming probes, treatment of that list was discontinued and intervention for a new list containing items belonging to a new semantic category initiated. If this criterion was not achieved after 8 consecutive days of intervention, treatment of a new list began to prevent participants from receiving treatment on a single list of words. Once participants advanced to a new treatment list, intervention for previously treated items ceased. This individualized nature of the treatment protocol resulted in some participants only receiving SFA treatment for 2 treatment lists (i.e., participants who never reached the criterion for advancement), with others receiving SFA treatment for 3 treatment lists (i.e. participants who showed rapid improvements on daily naming probes).

**Figure 2.**
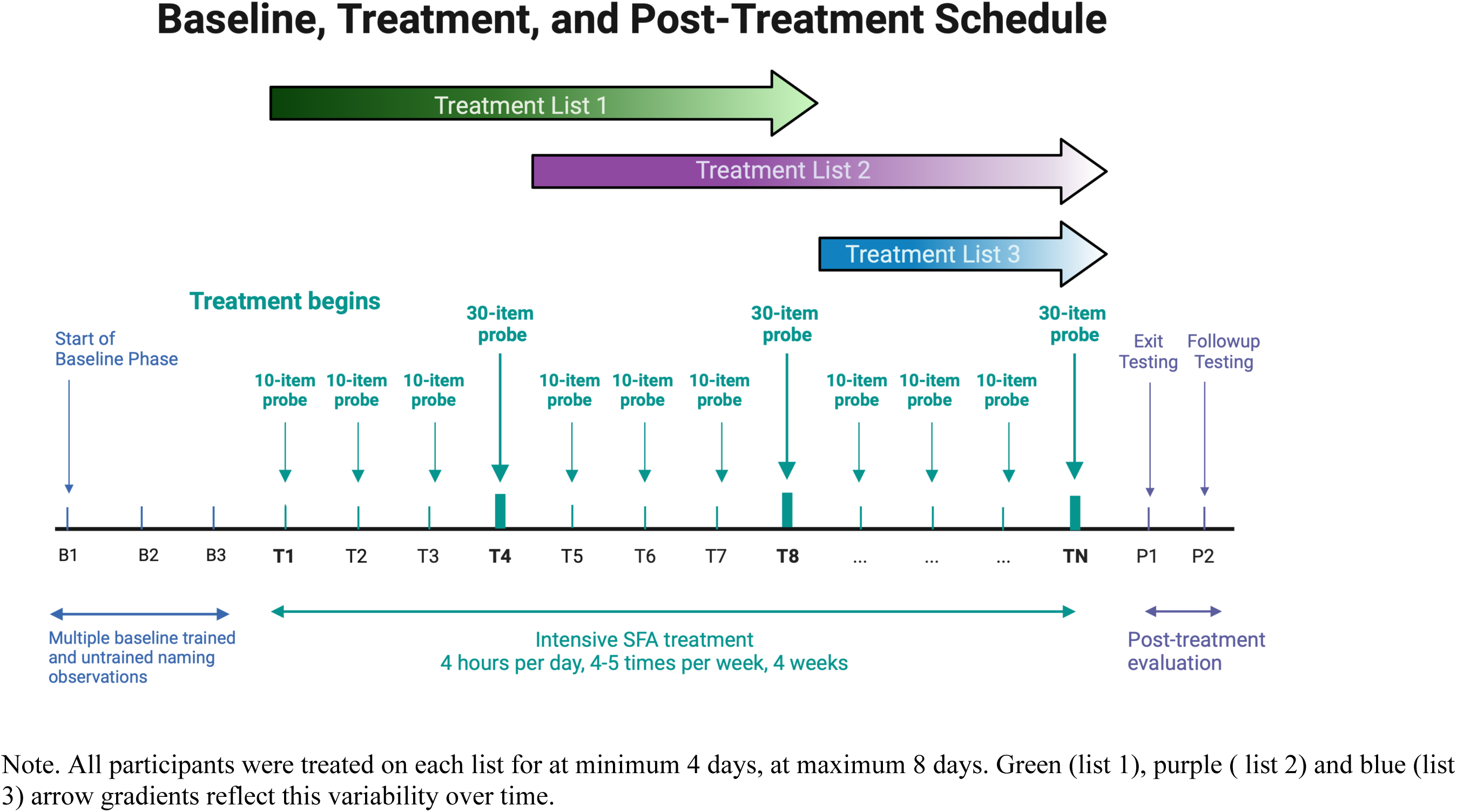
Depiction of study timeline including pre- and post-treatment evaluation and the daily probe naming schedule throughout SFA treatment.

Every 4 days, a master 30-item naming probe was collected in place of the daily 10-item probe. The 30-item probe contained trained (5 items) and untrained (5 items) target items for all 3 treatment lists (see Figure 2). Depending on how far into the 4-week intensive treatment participants were, this 30-item master naming probe allowed us to monitor naming accuracy for currently treated, yet-to-be treated, and previously treated items. For example, in the early phase of intervention, the 30-item probe allowed investigators to periodically continue probing baseline naming accuracy of yet-to-be treated words (e.g. from treatment lists 2 and 3, while list 1 was actively being treated). In the middle phase of intervention, when treatment of the first list was finished and treatment of a second list was occurring, the 30-item probe allowed investigators to periodically monitor how well individuals retained treatment effects of previously targeted words while continuing to gather pre-treatment baseline performance on yet-to-be treated words. In the later phase of intervention, when 1 or 2 lists had been treated, the 30-item probe allowed investigators to continue periodically monitoring retention of treatment effects for previously targeted words while treatment was still ongoing. This study design granted us the opportunity to monitor day-to-day changes in naming accuracy of actively treated words.

### Analyses

To estimate change in naming accuracy during daily naming probe performance, we used item-level generalized linear mixed-effects models. These models followed an established interrupted time series approach for multiple baseline designs (Huitema & Mckean, 2000). The models use generalized linear mixed effects models to test whether changes in performance across treatment phases (e.g. during treatment compared to baseline, or after treatment has been withdrawn) are statistically robust. They thus provide a statistically rigorous complement to effect-size-based tools like proportion of non-overlap or Tau-U, which have been criticized (Pustejovsky, 2019). ITS models have recently been applied to aphasia research to examine both unfolding aphasia treatment response and person-level predictors of such response (Evans, Cavanaugh, Quique, et al., 2021; Quique et al., 2022; Robinaugh et al., 2024; Sandberg et al., 2023; Swiderski et al., 2021). For a tutorial and additional information see Cavanaugh et al. (2023).

There are three key parameters in the standard interrupted time series model – (1) baseline slope, (2) level change, and (3) slope change. The baseline slope parameter models the average rate of change in item-level naming accuracy prior to treatment onset. The level change parameter is an estimate of the difference between performance on the first probe measured after the first unit of treatment predicted by baseline slope compared to performance on the first probe measured after the first unit of treatment predicted by the slope change. The slope change parameter models the average rate of change in performance during active intervention, relative to the trend estimated by the baseline slope parameter. The intercept estimate for our generalized linear mixed effects models represents naming performance of trained targets immediately before the first baseline observation. This interrupted time series approach was applied within a Bayesian framework, implemented in R via the brms package with a binomial probability distribution and a logistic link function (Bürkner, 2017, 2018, 2021) using ‘cmdstan’ (J. Gabry et al., 2024). For each model, the dependent variable was item-level naming performance (correct, incorrect). Model structures are presented in Supplemental Table 1.

The first aim of the study examined the shape of the learning curve for trained and untrained target items during intensive SFA and the influence of non-language cognitive functions on this learning (Aim 2). For Aim 1, population-level effects (i.e., fixed-effects in a frequentist framework) included simple effects of time series variables (baseline slope, level change, and slope change) and their interactions with word type (trained [reference], untrained but semantically related). Additionally, aphasia severity (z-transformed CAT mean T-scores) was included as a covariate to account for variability in language deficit severity among individuals in the study sample (Gravier et al., 2018). Group-level effects (i.e., random-effects in a frequentist framework) included intercepts and slopes for the key time series parameters by participant, and a group-level intercept for item. It is worth noting that treating list as a group-level effect, rather than a simple effect, entails that the model ignores effects of list order. This allowed us to handle unbalanced probe structures, multiple list and semantic categories, and the variable duration of treatment for each list within and across each participant. To address Aim 2, we evaluated the extent to which verbal recognition memory, visuospatial recall, and selective attention moderated treatment response as defined by time series variables (baseline slope, level change, and slope change) in separate, parallel models. Specifically, we added z-transformed cognitive scores as a third interaction term to the model used in Aim 1.

Prior distributions that supported our models were informed by our knowledge of the stimuli selection process and previous work using Bayesian statistical modeling in anomia treatments for aphasia (Boyle et al., 2023; Cavanaugh et al., 2022, 2023; Evans, Cavanaugh, Quique, et al., 2021). See Supplemental Material for additional information.

## Results

When estimating change in performance after level change, we used 4 treatment sessions, as this was the minimum number of treatment sessions that could have occurred before participants advanced to new lists and is the most conservative estimate. Model fixed effect estimates are discussed in terms of percent accuracy (i.e. percentage points), estimated from logit coefficients (see the markdown file included in Supplemental Material).

### Aim 1: Shape of Learning During Intensive SFA Intervention

Findings relevant to Aim 1 for both trained and untrained items are visualized in Figure 3. Aim 1 model output is located in Supplemental Table 2. The model-estimated average accuracy was 15.37% (95%CI [11.56%, 19.78%]) on treated items at the first baseline session.

**Figure 3.**
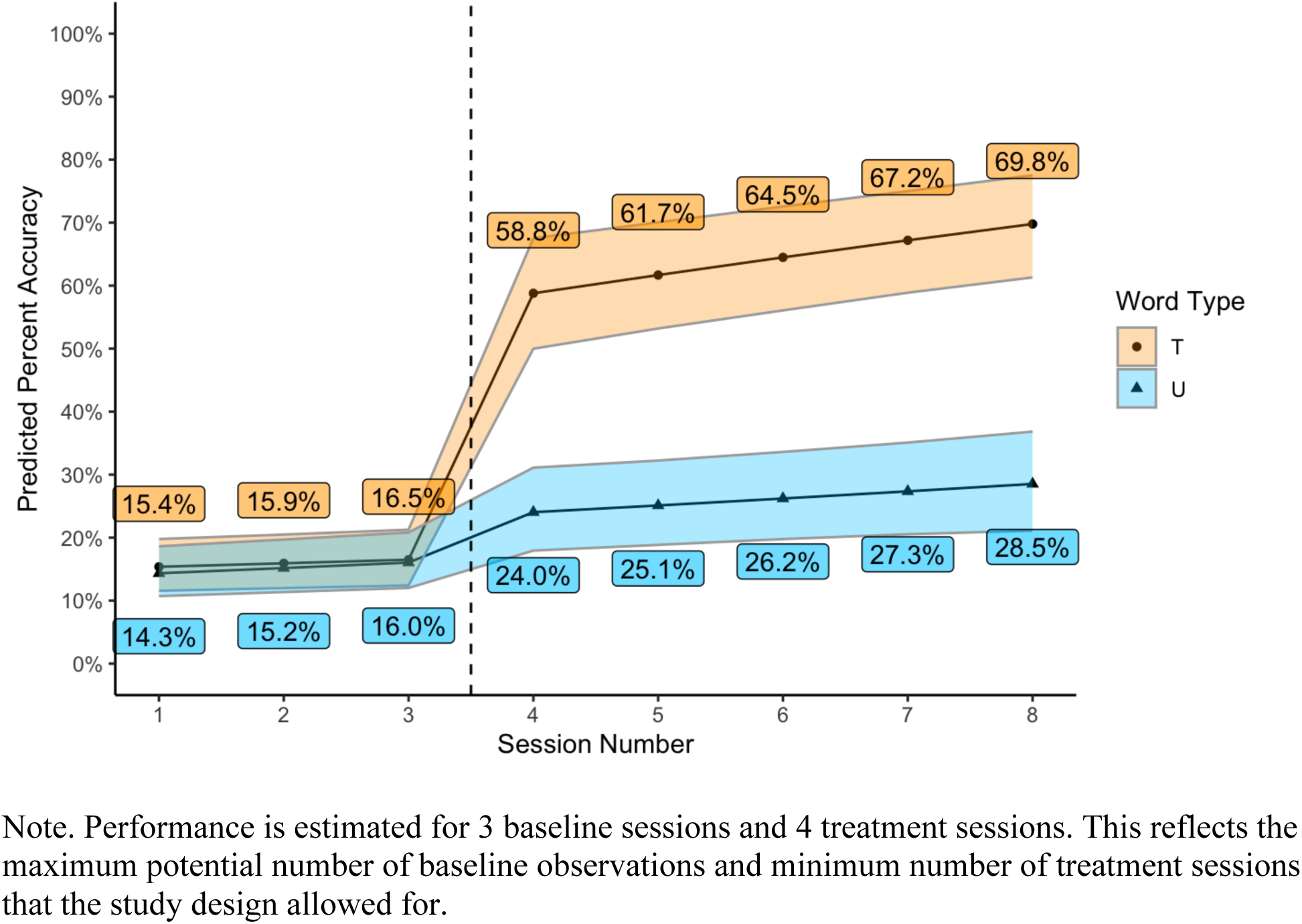
Rate of change in naming accuracy for trained (orange circle) and untrained (blue triangle) target items from the baseline to treatment phases (separated by dashed line).

For **trained** items, there was a small baseline trend which corresponded to an increase in estimated average naming accuracy from 15.4% at the first baseline session to 16.5% (Δ=1.1%, 95% CI[0.4%, 1.9%]) at the final baseline session. There was a large positive effect of level change, which was associated with an average increase from 16.5% to 58.8% (Δ=42.3%, 95% CI[35.9%, 48.3%]) on treated items following the first 4 hours of intensive SFA treatment. There was a slightly positive effect of slope change (the rate of change in naming accuracy during treatment relative to baseline rate) for trained items. Model-estimated accuracy for naming of trained items increased from 61.7% to 69.8% (Δ=8.1%, 95% CI[4.7%, 11.6%]) after four days of intensive SFA.

When comparing naming accuracy of **untrained** but semantically related items to trained items, there was evidence of a positive interaction of word type and baseline slope. Although the 95% credible interval (CI) of this effect overlapped 0 (Supplemental Table 2), 97.8% of the posterior distribution was greater than 0, suggesting a high probability the effect was greater than 0. The average estimated naming accuracy of untrained items increased from 14.3% at the first baseline session to 16.0% (Δ=1.7%, 95% CI[0.1%, 25.5%]) at the final baseline session. There was a negative interaction between level change and word type consistent with a smaller positive effect of the introduction of treatment on untreated but semantically related items than on trained items. Average accuracy on untreated items increased from 16.0% to 24.0% (Δ=8.0%, 95% CI[3.8%, 12.6%] after the first 4 hours of intensive SFA treatment. There was also a negative interaction between slope change and word type. The rate of change in naming accuracy during treatment for untrained items was less positive than for trained items, and essentially the same as the small positive rate of change during baseline for untrained items. By the end of the 4^th^ treatment session, the estimated average naming accuracy on untrained items increased from 25.1% to 28.5% (Δ=3.4%, 95% CI[0.0%, 6.7%], PD=100%).

### Aim 2: Influence of Cognitive Function on Shape of Learning During Intensive SFA Intervention

#### Verbal Recognition Memory

Supplemental Table 3 includes model output associated with verbal recognition memory, and Supplemental Table 4 presents estimated naming accuracy of trained and untrained items across baseline, level change, and treatment slope phases for individuals with verbal recognition memory scores that fell 1 standard deviation or greater above the sample mean (hereafter “above average”) and for individuals with verbal recognition memory scores that fell 1 standard deviation or greater below the sample mean (hereafter “below average”).

For **trained** items, there was a positive two-way interaction between level change and verbal recognition memory score. The 95% CI of this effect overlaps 0, however 96.6% of the posterior distribution was greater than 0, suggesting a high probability the effect was greater than 0. Participants with above average verbal recognition memory scores increased their naming accuracy of trained items from 17.6% to 66.4% (Δ=48.8%, 95% CI[41.2%, 55.7%]) after the first four hours of treatment. In comparison, participants with below average verbal recognition memory scores increased their naming accuracy from 15.6% to 50.2% (Δ=34.6%, 95% CI[25.8%, 43.2%]). There was weak (probability of direction [PD]=91.1%) evidence for a small effect of the two-way interaction between verbal recognition memory and slope change, which favored participants with above average verbal recognition memory abilities.

When examining the influence of verbal recognition memory on the difference in naming accuracy for **untrained** compared to trained items across the three time series variables, evidence supporting all three-way interaction effects was weak (all PDs < 74%) and the directions of each interaction effect were variable (see model outputs in supplemental material).

#### Visuospatial Memory

Supplemental Table 5 includes model output associated with visuospatial memory, and Supplemental Table 6 presents estimated naming accuracy of trained and untrained items across baseline, level change, and slope change phases for individuals with above average visuospatial recall scores and for individuals with below average visuospatial recall scores.

For **trained** items, there was a positive two-way interaction between visuospatial recall score and baseline slope. The rate of change in naming accuracy across baseline observations for trained items was marginally more positive for participants with above average visuospatial recall scores compared to participants with below average scores. The 95% CI of this effect overlaps with 0 (see Supplemental Table 5), however 95.7% of the posterior distribution was greater than 0. There was a strong and positive two-way interaction between visuospatial recall score and level change. Participants with above average visuospatial recall scores increased their naming accuracy of trained items from 15.4% to 66.2% (Δ=50.8%, 95% CI[42.7%, 58.1%]) at the level change. Comparatively, participants with below average visuospatial recall scores increased from 17.7% to 50.3% (Δ=32.6%, 95% CI[24.2%, 40.8%]) at the level change. There was no evidence supporting a two-way interaction between visuospatial recall and slope change (PD = 59.7%).

The three-way interaction between **untrained** targets compared to trained targets for participants with above and below average visuospatial recall scores revealed a small negative trend (PD=98.6%) in the baseline slope, suggesting that individuals with above average visuospatial memory scores improved more on treated than untreated words during the baseline phase. The three-way interaction between level change, word type, and visuospatial recall yielded a negative effect (PD=96.6%) This suggests that participants with below average visuospatial recall scores experienced more comparable level change effects for trained and untrained items, whereas individuals with above average visuospatial recall scores showed much larger level change effect for trained compared to untrained items. There was no evidence supporting the three-way interaction between slope change, word type, and visuospatial recall score, and the direction of the effect was unclear (PD=66.5%).

#### Sustained Attention

Supplemental Table 7 includes model output associated with visuospatial memory, and Supplemental Table 8 presents estimated naming accuracy of trained and untrained items across baseline, level change, and treatment slope phases for individuals with above average sustained attention scores and for individuals with below average sustained attention scores.

For **trained** items, there was no evidence for two-way interactions between sustained attention score and baseline slope (PD = 79.9%) or level change (PD = 69.9%). There was weak evidence supporting a two-way interaction between sustained attention score and slope change (PD=98.0%), suggesting that rate of change in percentage points for trained items was slightly more positive for those with above-average sustained attention scores than those with below-average scores. There were no reliable 3-way interactions for sustained attention (all PDs <78%; see model outputs in the supplemental R-Markdown files).

## Discussion

In this study, we aimed to model the shape of learning that occurs during the time course of treatment while individuals with post-stroke aphasia engage in SFA (Aim 1), an anomia intervention that is commonly used in both clinical practice and research (Efstratiadou et al., 2018; Raymer & Roitsch, 2023b). Additionally, we aimed to determine which cognitive functions were associated with learning during treatment (Aim 2).

### Study Aim 1: Shape of Learning During Aphasia Treatment

The present study found: (1) a small increase in naming accuracy across baseline observations (i.e. baseline slope) which was greater for untrained than trained target words; (2) a large, positive increase in naming accuracy after the first four hours of SFA treatment (i.e. at the level change) which was greater for trained than untrained words; and (3) a slightly positive change in naming accuracy across treatment sessions compared to baseline (i.e. slope change), which was again greater for trained than untrained target words.

The rising baseline slope reported in this investigation is consistent with findings from other (limited) studies that have used interrupted time series models to examine aphasia treatment response (Evans, Cavanaugh, Quique, et al., 2021; Swiderski et al., 2021). Specifically, individuals with aphasia demonstrate slight improvements in production of trained and untrained target items from one baseline session to the next, in the absence of direct intervention. The rising baseline slope for trained and untrained target items could represent an effect of repeated exposure, where repeatedly viewing a stimulus over the course of 1-2 days facilitates slightly increased likelihood for successful retrieval (Creet et al., 2019). However, it is worth remarking that the improvements made throughout the baseline phase were clinically unremarkable, translating to total increases of 2% or less across the baseline phase for trained and untrained targets.

Among the three interrupted time series variables in our model, the effect of level change yielded the largest percentage point change. On average, individuals in our sample attained the majority of SFA-related naming improvements early into treatment, after the first four hours. We also found a positive though small effect of slope change for trained target items, indicating that participants continued to improve incrementally in naming trained items from session to session at a faster rate during treatment than during the baseline phase. Taken together, the level change and slope change findings are consistent with our hypotheses. We anticipated that, much like results by Dignam et al. (2023) and Simic et al. (2020), individuals with aphasia would exhibit rapidly emerging SFA treatment gains, with most of the benefit of SFA intervention being observed after the first 4 hours of SFA. These results corroborate the importance of early treatment response as a potential indicator of an individual’s potential to benefit from an intervention (Dignam et al., 2023).

The relative magnitude of the level change and slope change results observed is also consistent with findings from the small number of studies that have used interrupted time series models to examine aphasia treatment response (Evans, Cavanaugh, Quique, et al., 2021; Sandberg et al., 2023; Swiderski et al., 2021). Swiderski and colleagues (2021) used interrupted time series models in a meta-analysis of single-subject design studies examining treatment effects for Treatment of Underlying Forms (TUF), a widely-researched sentence-level treatment (Thompson & Shapiro, 2005). They reported a level change effect for trained stimuli (β=2.46) that was much greater than the slope change effect (β=0.51). Evans et al. (2021) paired anomia treatment (semantic feature verification) with an intervention aimed at targeting speed accuracy tradeoffs and used interrupted time series models to gauge the effect of their novel treatment pairing on naming accuracy of trained and untrained targets. The investigators reported a large level change effect for trained items (β=0.56) with a positive yet modest slope change effect (β=.03). Sandberg et al. (2023) used interrupted time series models in a meta-analysis of single-subject design studies of another semantically-focused anomia treatment, Abstract Semantic Associative Network Training (AbSANT: Sandberg & Gray, 2020). They also reported large level change effects for trained targets (β=.80) and a positive but modest slope change effect (β=.03).

These studies using interrupted time series models to examine aphasia treatment efficacy consistently report an early and substantial effect of treatment - one that is captured via the level change parameter. This suggests that across anomia and sentence-level interventions, most of the re-learning in aphasia treatment tends to occur within the first few hours of treatment. ITS models directly capture this feature of aphasia treatment response, drawing attention to a pattern that has previously gone unnoticed in the majority of group aphasia treatment studies, in part because they have tended to focus on comparisons of pre- and post-treatment behaviors using effect size measures (Beeson & Robey, 2006). This early phase of treatment has also been associated with longer-term maintenance of therapy gains (Dignam et al., 2023; Simic et al., 2020). Thus, early improvement in aphasia treatment is a person-specific factor that may be used to predict patient outcomes and inform rehabilitation planning.

In comparison to trained items, participants in our sample demonstrated less robust improvements in naming accuracy for untrained items, both at the level change and throughout treatment (i.e., smaller slope change effects). We anticipated that treatment-driven improvements would be more positive for trained items; thus, this result is generally consistent with our hypothesis. The percentage point increases in naming accuracy of semantically-related untrained target words at the level change do not provide strong evidence of generalization of treatment effects over the course of intensive SFA. Other studies investigating generalization of treatment effects to untrained target words among individuals with aphasia have also reported limited generalization to untrained stimuli (Nickels, 2002; Wisenburn & Mahoney, 2009; Estratiadou et al., 2018). Our findings are hence consistent with this pattern of relatively weak response generalization for anomia treatment, i.e., limited treatment-related changes for untrained targets.

However, the current findings do suggest that the relatively small changes detected for untreated semantically-related words are likely due to SFA treatment, and not simply the opportunity to name the untreated items repeatedly. Creet et al. (2019) showed that repeated naming attempts may result in positive changes in word retrieval for at least some people with aphasia, and Gravier et al. (2018), Rider et al. (2008), and Wambaugh et al. (2013) speculated that such exposure effects may be behind apparent response generalization effects for SFA. However, the current findings revealed that the majority of improvements made in naming of untrained semantically-related items occurred at the level change, rather than being distributed evenly across the course of treatment, as would be expected if these improvements were due to exposure. Instead, gains in naming accuracy for untrained items were largest in magnitude after the first four hours of SFA treatment, compared to slight changes noted during baseline and later treatment sessions. These findings indicate that the shape of SFA response generalization mirrors the shape of learning for trained targets, but with smaller-magnitude changes.

### Aim 2: Cognitive Predictors of Learning During Aphasia Treatment

Aphasia rehabilitation is increasingly viewed as a re-learning process (Dignam et al., 2016; Helm-Estabrooks, 2002; Nunn et al., 2023; Peñaloza et al., 2022), and an individual’s potential for learning may serve as a person-specific predictor of rehabilitation outcomes (Vallila-Rohter, 2017). Learning is a complex cognitive capacity that is interdependent on multiple domain specific cognitive functions, including memory and attention. To date, multiple prior studies have reported that relatively stronger verbal memory, visuospatial recall, and sustained attention scores are associated with robust aphasia treatment outcomes (Diedrichs et al., 2022; Dignam et al., 2017; Gilmore et al., 2019; Lambon Ralph et al., 2010; Seniów et al., 2009). These empirical studies have examined therapy gains by comparing pre- and post-treatment language outcomes, without characterizing the learning process that occurs as treatment unfolds.

In the current study, verbal recognition memory and visuospatial recall both emerged as moderators of changes in trained word naming accuracy over the treatment time series.

Specifically, above-average verbal recognition memory and visuospatial recall abilities were associated with larger increases in trained naming accuracy at the first treatment probe (Aim 1). This suggests individuals with relatively strong memory abilities are advantaged to benefit early in the course of treatment, a crucial phase that may be predictive of long-term outcomes (Dignam et al., 2023; Simic et al., 2020). Verbal short-term memory and visuospatial recall serving as significant moderators of the magnitude of improvements for trained items at the level change is consistent with findings from studies that compared pre- and post-treatment naming performance (Diedrichs et al., 2022; Dignam et al., 2017; Gilmore et al., 2019; Goldenberg et al., 1994; Lambon Ralph et al., 2010), suggesting these cognitive functions as potential pillars supporting aphasia treatment success, including incremental learning during aphasia treatment.

The memory measures that were predictive of anomia treatment outcomes, both in the current study and previous studies, merit further discussion. We measured verbal recognition memory via the Paired Associate Learning subtest of the Camden Memory Test, which requires test-takers to associate pairs of unrelated words presented orthographically, broadly gauging ability to encode and immediately retrieve verbal information. Previous studies have also found performance on the Camden Memory Test to be predictive of anomia treatment outcomes (e.g., Gilmore et al., 2019; Lambon Ralph et al., 2010). Performance in this verbal paired-associate learning task benefits from having relatively rich semantic representations that can be exploited for successfully associating unrelated words and recalling that ad-hoc semantic association. This feature of this paired-associate task offers a natural explanation for why individuals with aphasia who are relatively good at the verbal paired associate task are more likely to benefit from SFA, a semantically-oriented intervention that leverages lexical semantic knowledge and connections. Of note, the paired-associate learning task only measures short-term encoding of information without placing demands on long-term recall, another crucial cognitive capacity supportive of learning potential. Thus, our findings in combination with others (Dignam et al., 2016), underline the importance of **short-term** verbal encoding as a predictor of early anomia treatment response, leaving the role of long-term memory abilities largely unexplored.

The relationship between visuospatial recall (measured using immediate recall of the Rey Complex Figure) and aphasia rehabilitation outcomes may not be as straightforward or obvious as the connection to verbal recognition memory. Several previous investigations have also identified Rey Complex Figure Copy performance as a predictor of aphasia treatment outcomes (Gilmore et al., 2019; Goldenberg et al., 1994; Lambon Ralph et al., 2010), while others have not (Conroy & Scowcroft, 2012; Diedrichs et al., 2022; Fillingham et al., 2006; Rose et al., 2013; Votruba et al., 2013). An inherent advantage to characterizing the relationship between visuospatial recall and aphasia treatment is its *lack* of reliance on linguistic processing: this feature of these tasks may be especially advantageous for examining the association of memory capacity with treatment outcomes in the absence of language-processing demands that may be problematic in a language impaired population (Dignam et al., 2017; Goldenberg et al., 1994).

Beyond measuring memory capacity using non-linguistic means, visuospatial processing and recall abilities may be important for the organization of semantic or conceptual representations. For example, Viganò et al. (2021) found that healthy individuals activated brain regions that were used during visuospatial tasks (i.e. entorhinal cortex within hippocampus) while making semantic judgments about recently learned novel words. These novel words varied along two dimensions of semantic representation (pitch and size), and the investigators reported that the organization of such semantic representations in relevant sensory cortices and entorhinal cortex had similar grid-like structure to visuospatial representations. Viganò et al.’s findings suggest that visuospatial and semantic conceptual representations may be supported by the same neurocognitive systems. If so, individuals with better visuospatial processing and memory might also be expected to have better conceptual processing (Bottini & Doeller, 2020; Constantinescu et al., 2016; Epstein et al., 2017; Viganò et al., 2021). In addition, Diedrichs et al. (2022) discuss how visuospatial recall fundamentally supports higher order cognitive abilities such as executive functioning that are important for learning and have been associated with aphasia treatment outcomes (Simic et al., 2019), which may help explain its influence in the context of the present findings.

There was less clear evidence regarding which non-language cognitive factors influenced changes in naming of untrained target words (i.e. response generalization) over the course of SFA treatment. Evidence for the influence of verbal recognition memory and sustained attention on untrained-item changes was weak. However, there was a small effect of visuospatial recall such that individuals in our sample with relatively stronger visuospatial recall performance were more likely to generalize treatment effects to untrained items at the level change - the point of greatest treatment-related change in naming performance. Response generalization for semantically-based anomia treatment is hypothesized to involve successful activation of relevant semantic/conceptual representations (Boyle, 2010; Foygel & Dell, 2000; Kiran & Bassetto, 2008). The positive relationship between visuospatial recall and SFA-related changes for untrained items may be because visuospatial and semantic/conceptual processing appear to be related, as noted above (Viganò et al., 2021). Individuals with better visuospatial processing and recall may have better access to or better organization of semantic/conceptual representations, supporting response generalization to untrained but semantically-related items. This possibility would be consistent with the account suggested above for the positive effect of visuospatial processing ability on changes for trained items. Together with the inconsistent previous findings regarding the influence of visuospatial recall on aphasia treatment outcomes (Gilmore et al., 2019; Goldenberg et al., 1994; Lambon Ralph et al., 2010); (Conroy & Scowcroft, 2012; Diedrichs et al., 2022; Fillingham et al., 2006; Rose et al., 2013; Votruba et al., 2013), the current findings underline the need for further work aimed at uncovering the cognitive mechanisms behind this interesting relationship.

The lack of influence of attention scores on the unfolding treatment response, both for trained and untrained target items, is consistent with several other findings (Yeung & Law, 2010). Although sustained attention is crucial for focus in the therapy room, perhaps it is less pivotal in supporting learning capacity than the memory sub-domains that yielded significant findings in our investigation. The robust contributions of memory abilities to SFA treatment response found here ground these effects in explicit learning of declarative (e.g. verbal) content, which resembles learning that occurs in language treatment sessions over the course of anomia rehabilitation. Memory and learning require successful encoding, consolidation, and retrieval of learned information. Disruptions at any of these crucial learning stages may lead to suboptimal treatment response. The memory measures used in this study, as well as in the majority of other studies examining cognitive predictors of aphasia treatment outcomes, use immediate recall to gauge verbal or visuospatial memory capacities. These measures likely inform us about how well the individuals in our samples can encode and temporarily store information, but they do not offer insight into how well the information becomes consolidated and retrieved long-term (across days, weeks, or months). To fully understand the learning process that occurs during anomia treatment, which could provide further insight into the neural and cognitive systems that support aphasia treatment response, these underexplored components of learning must be systematically investigated.

### Clinical Implications

Variability in aphasia treatment response presents a barrier to the clinical management of post-stroke aphasia. Accounting for this individual variability is key to delivering precise intervention to people with aphasia, and integral to optimizing language therapy outcomes. Results related to the first study aim revealed that most gains in trained and untrained naming accuracy occurred early in the course of treatment, after just the first four hours of SFA. The clinical significance of the robust level change effect is best contextualized when also considering results reported by Simic et al. (2020) and Dignam et al. (2023). Both Simic et al. and Dignam et al. reported that early treatment response was predictive of ultimate treatment outcomes, such that patients who demonstrated rapid response to treatment made the most improvements by the end of intervention and later (at one month follow up). Treatment-related changes within the first several hours of intervention also represent the largest proportion of naming improvement across the time course of treatment, and they may serve as a person-specific prognostic indicator for aphasia therapy success (Dignam et al., 2023). Given this, it will be beneficial for speech language pathologists to monitor early SFA treatment response to gauge the prognosis for treatment outcomes and modify their treatment plans accordingly: limited response following those initial hours of treatment may indicate that both additional gains and ultimate outcomes are likely to be limited, suggesting a change in treatment approach may be warranted. Although it appears from our findings that the majority of treatment gains are made within the first four hours of SFA intervention, this does not suggest that therapy should be delivered for only that amount of time. After the robust level change, individuals continued to demonstrate modest increases in their naming accuracy of trained target words from one treatment session to the next. Thus, intervention for people with aphasia (both SFA and other treatments) requires time, repetition, practice, and individualization.

Consistent with findings from multiple other studies (Dignam et al., 2016, 2017; Gilmore et al., 2019; Lambon Ralph et al., 2010), our results implicate memory function as a key cognitive pillar supporting word relearning during anomia treatment. A benefit of determining which cognitive factors predict outcomes for particular aphasia treatments is that those cognitive abilities can be evaluated at the pre-treatment diagnostic phase of aphasia service delivery. Such pre-treatment evaluations can allow clinicians to efficiently determine poor and strong “fits” for specific treatment protocols before treatment is initiated, optimizing use of limited therapy visits covered by insurance companies. Clinicians may consider including measures of linguistic and non-linguistic memory and learning into their diagnostic batteries to inform treatment planning.

### Limitations

Participants advanced from one treatment list to the next at different rates, based on their daily naming probe performance. Although individualized treatment lists and variable time spent training those lists serve as strengths highlighting the importance of tailoring anomia treatment to the individual, they also pose as challenges in analyzing these data at the group level. Findings presented here represent group averages that may overlook inter-individual variability over the course of treatment. Treatment duration in this investigation was long (2 hours per session), which is not representative of duration of language therapy sessions in clinical practice. Additionally, all participants in our sample were recruited at the chronic phases of their aphasia recovery trajectories, to control for potential effects of spontaneous recovery on treatment-related changes in performance. It is therefore unclear whether and how our findings generalize to the acute and subacute phases of recovery.

## Conclusions

The current study carefully examined incremental learning during aphasia treatment to dissect the unfolding SFA treatment response. The findings revealed that the bulk of treatment gains come early in the time course, even more so for individuals with cognitive strengths in verbal recognition memory. These findings can inform current clinical practice for anomia treatment, and they point to the important but overlooked role of learning-related processes and mechanisms in aphasia treatment, paying the groundwork for future investigations.

## Data Availability

Because of information security requirements associated with the project, direct data sharing will not be possible. However, we encourage individual investigators to contact us.

## Acknowledgments

Alyssa Autenreith, Rebecca Ruffing, Emily DeFrancesco, and Angela Grzyzbowski served as the treating clinicians for the parent study, collected pre- and post-treatment evaluation data from participants, and performed key administrative tasks to enable successful data collection and analysis that supported this investigation. We are deeply grateful for their contributions to this work. We are also immensely appreciative of the individuals with aphasia who dedicated their time and effort to participating in the study.

## Supplemental Material

**Supplemental Table 1.**
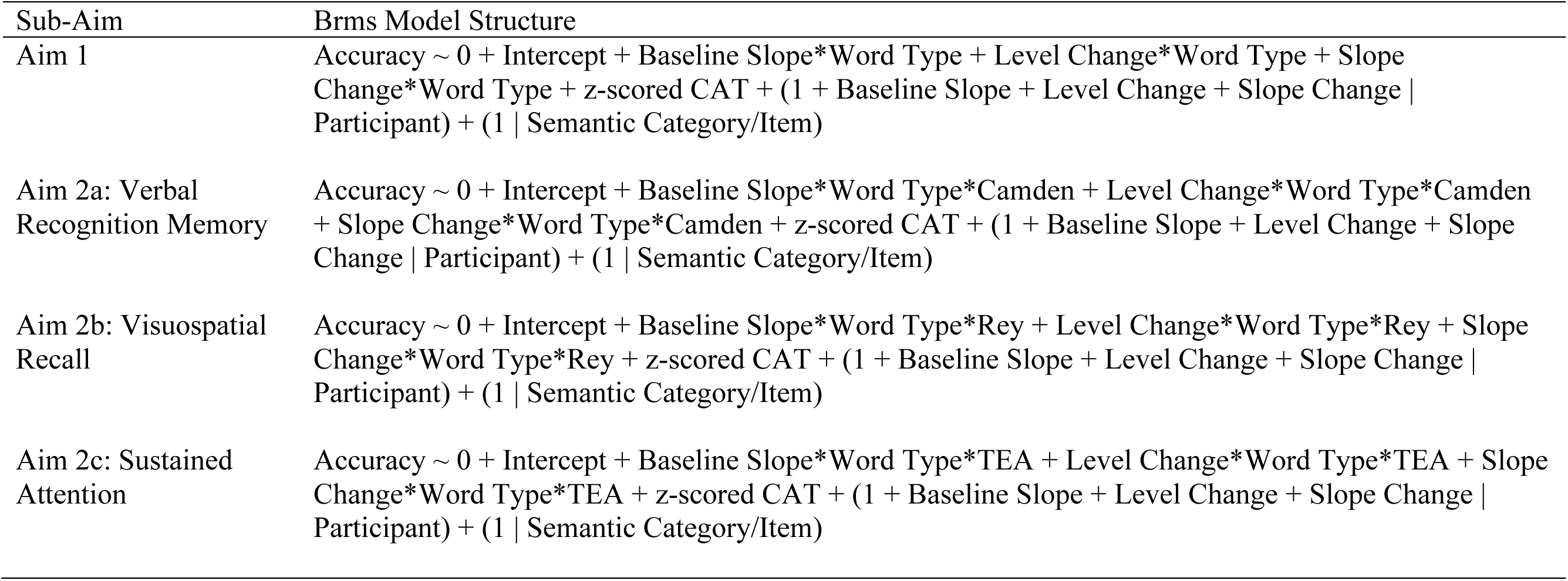
Model structures used to address all study aims via the brms package in R Studio.

**Supplemental Table 2.** Aim 1 model coefficients.

| Coefficient | Estimate | Est. Error | Lower 95% CI | Upper 95% CI | Rhat | Bulk ESS | Tail ESS |
| --- | --- | --- | --- | --- | --- | --- | --- |
| Intercept | -1.76 | 0.16 | -2.08 | -1.44 | 1.00 | 3307 | 4387 |
| Baseline Slope | 0.04 | 0.01 | 0.02 | 0.07 | 1.00 | 5923 | 5933 |
| Type (U) | -0.11 | 0.11 | -0.32 | 0.11 | 1.00 | 9838 | 6056 |
| Level Change | -1.95 | 0.13 | 1.69 | 2.21 | 1.00 | 6561 | 6678 |
| Slope Change | 0.08 | 0.03 | 0.02 | 0.14 | 1.00 | 7495 | 6300 |
| Z-scored CAT | 0.68 | 0.14 | 0.42 | 0.98 | 1.00 | 2425 | 3330 |
| Baseline Slope:Type (U) | 0.02 | 0.01 | 0.00 | 0.05 | 1.00 | 10388 | 6004 |
| Level Change: Type (U) | -1.51 | 0.14 | -1.78 | -1.24 | 1.00 | 11139 | 6809 |
| Slope Change: Type (U) | -0.09 | 0.03 | -0.15 | -0.03 | 1.00 | 11468 | 7028 |

*Note: The intercept is interpreted for the reference category of Type, which is trained target words. (U) = untrained but semantically-related target words. Model coefficients are presented in logits.*

**Supplemental Table 3.** Aim 2 model coefficients with z-transformed Camden Verbal Memory score as the cognitive variable of interest.

| Coefficient | Estimate | Est. Error | Lower 95% CI | Upper 95% CI | Rhat | Bulk ESS | Tail ESS |
| --- | --- | --- | --- | --- | --- | --- | --- |
| Intercept | -1.76 | 0.16 | -2.08 | -1.44 | 1.00 | 3198 | 4892 |
| Baseline Slope | 0.04 | 0.01 | 0.02 | 0.07 | 1.00 | 5576 | 5411 |
| Type (U) | -0.11 | 0.11 | -0.32 | 0.11 | 1.00 | 10106 | 6398 |
| Z-scored Camden | 0.01 | 0.14 | -0.28 | 0.29 | 1.00 | 3829 | 4791 |
| Level Change | 1.94 | 0.13 | 1.69 | 2.19 | 1.00 | 6463 | 6099 |
| Slope Change | 0.08 | 0.03 | 0.03 | 0.14 | 1.00 | 7083 | 6114 |
| Z-scored CAT | 0.67 | 0.14 | 0.41 | 0.96 | 1.00 | 2422 | 4174 |
| Baseline Slope:Type (U) | 0.02 | 0.01 | 0.00 | 0.05 | 1.00 | 10454 | 6317 |
| Baseline Slope: Z-scored Camden | 0.02 | 0.01 | -0.01 | 0.05 | 1.00 | 5430 | 5508 |
| Type (U): Z-scored Camden | 0.09 | 0.11 | -0.12 | 0.30 | 1.00 | 10657 | 6380 |
| Type (U): Level Change | -1.49 | 0.14 | -1.76 | -1.22 | 1.00 | 10041 | 6396 |
| Z-scored Camden: Level Change | 0.25 | 0.13 | -0.02 | 0.51 | 1.00 | 6542 | 5663 |
| Type (U): Slope Change | -0.09 | 0.03 | -0.15 | -0.03 | 1.00 | 10967 | 6574 |
| Z-scored Camden: Slope Change | 0.04 | 0.03 | -0.02 | 0.10 | 1.00 | 6940 | 7022 |
| Baseline Slope: Type (U): Z-scored Camden | -0.01 | 0.01 | -0.03 | 0.02 | 1.00 | 9328 | 6623 |
| Level Change: Type (U): Z-scored Camden | -0.08 | 0.15 | -0.37 | 0.21 | 1.00 | 9603 | 6477 |
| Slope Change: Type (U): Z-scored Camden | -0.02 | 0.03 | -0.09 | 0.05 | 1.00 | 9062 | 5655 |

**Supplemental Table 4.**
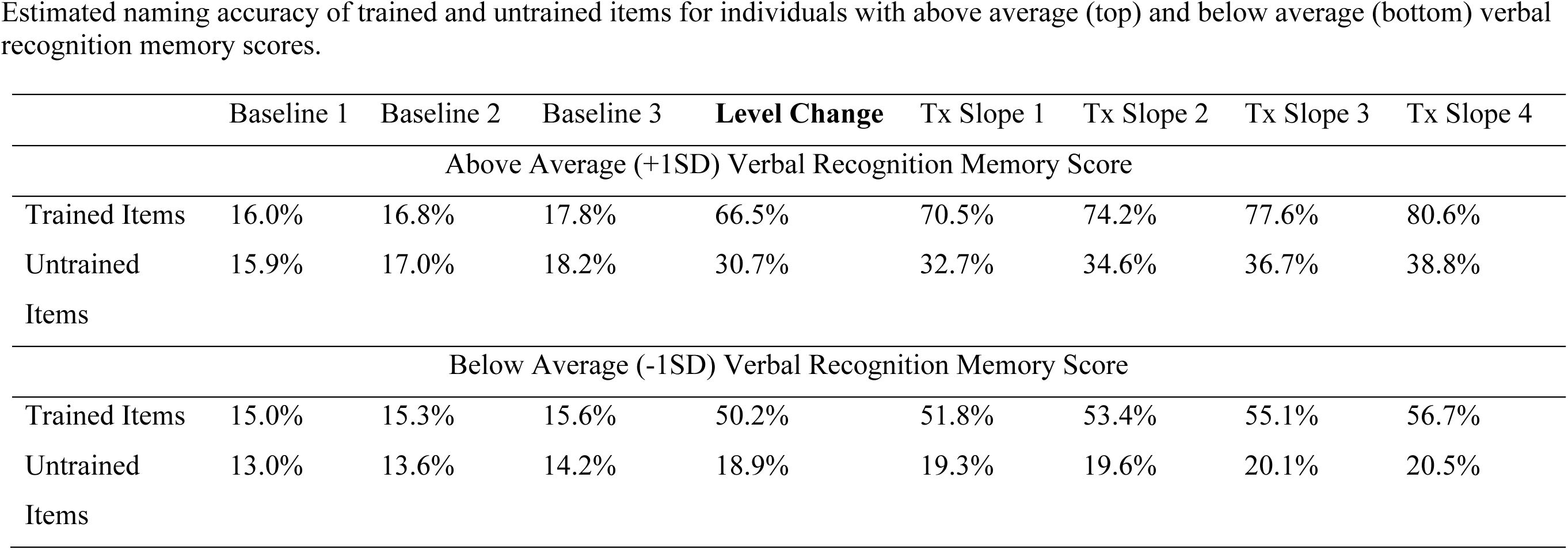
Estimated naming accuracy of trained and untrained items for individuals with above average (top) and below average (bottom) verbal recognition memory scores.

**Supplemental Table 5.**
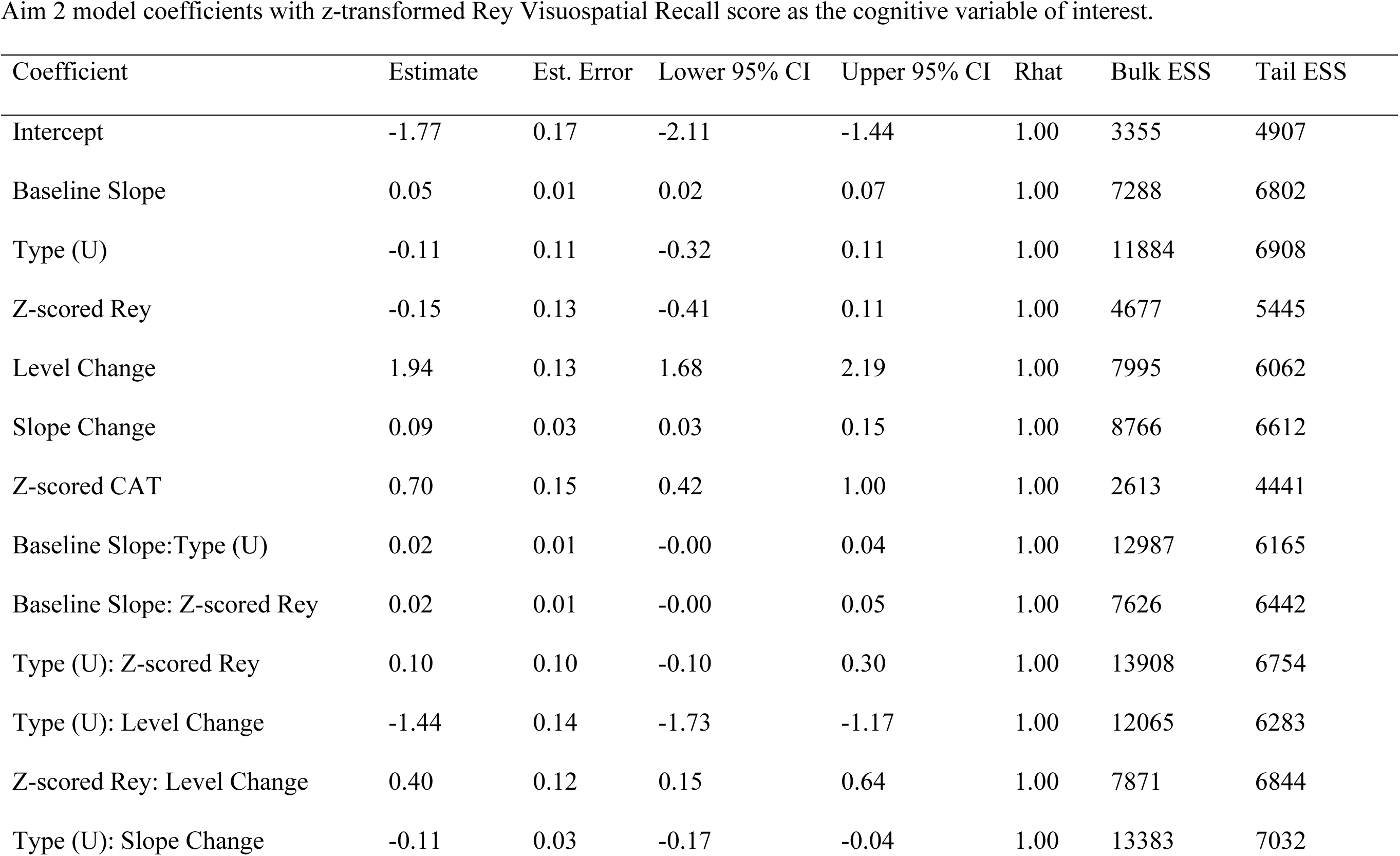
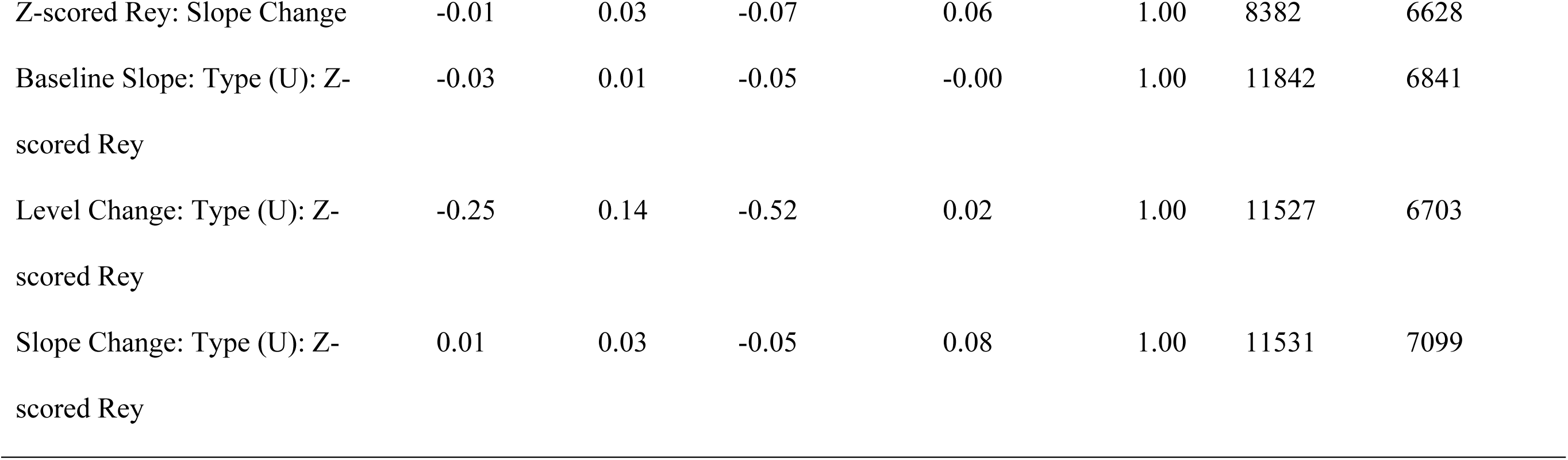
Aim 2 model coefficients with z-transformed Rey Visuospatial Recall score as the cognitive variable of interest.

**Supplemental Table 6.** Estimated naming accuracy of trained and untrained items for individuals with above average (top) and below average (bottom) visuospatial recall scores.

|  | Baseline 1 | Baseline 2 | Baseline 3 | Level Change | Tx Slope 1 | Tx Slope 2 | Tx Slope 3 | Tx Slope 4 |
| --- | --- | --- | --- | --- | --- | --- | --- | --- |
| Above Average (+1SD) Visuospatial Recall Score |  |  |  |  |  |  |  |  |
| Trained Items | 13.7% | 14.5% | 15.3% | 66.2% | 69.4% | 72.4% | 75.2% | 77.8% |
| Untrained Items | 13.5% | 14.2% | 15.0% | 26.2% | 27.2% | 28.2% | 29.3% | 30.3% |
| Below Average (-1SD) Visuospatial Recall Score |  |  |  |  |  |  |  |  |
| Trained Items | 17.2% | 17.5% | 17.8% | 50.5% | 53.4% | 56.3% | 59.2% | 62.0% |
| Untrained Items | 15.0% | 15.8% | 16.8% | 23.3% | 24.1% | 24.9% | 25.8% | 26.7% |

**Supplemental Table 7.** Aim 2 model coefficients with z-transformed TEA score as the cognitive variable of interest.

| Coefficient | Estimate | Est. Error | Lower 95% CI | Upper 95% CI | Rhat | Bulk ESS | Tail ESS |
| --- | --- | --- | --- | --- | --- | --- | --- |
| Intercept | -1.76 | 0.16 | -2.08 | -1.44 | 1.00 | 2621 | 4669 |
| Baseline Slope | 0.04 | 0.01 | 0.01 | 0.07 | 1.00 | 5048 | 5447 |
| Type (U) | -0.11 | 0.11 | -0.33 | 0.10 | 1.00 | 7694 | 6700 |
| Z-scored TEA | 0.02 | 0.14 | -0.24 | 0.29 | 1.00 | 3513 | 4788 |
| Level Change | 1.96 | 0.13 | 1.71 | 2.22 | 1.00 | 5162 | 5813 |
| Slope Change | 0.08 | 0.03 | 0.02 | 0.13 | 1.00 | 6140 | 6301 |
| Z-scored CAT | 0.66 | 0.14 | 0.40 | 0.95 | 1.00 | 2113 | 3727 |
| Baseline Slope:Type (U) | 0.03 | 0.01 | 0.00 | 0.05 | 1.00 | 8257 | 6226 |
| Baseline Slope: Z-scored TEA | -0.01 | 0.01 | -0.04 | 0.02 | 1.00 | 4431 | 4744 |
| Type (U): Z-scored TEA | 0.12 | 0.11 | -0.08 | 0.33 | 1.00 | 8146 | 6066 |
| Type (U): Level Change | -1.54 | 0.14 | -1.81 | -1.27 | 1.00 | 8332 | 7022 |
| Z-scored TEA: Level Change | 0.07 | 0.13 | -0.19 | 0.34 | 1.00 | 5187 | 5450 |
| Type (U): Slope Change | -0.09 | 0.03 | -0.15 | -0.02 | 1.00 | 8401 | 6784 |
| Z-scored TEA: Slope Change | 0.06 | 0.03 | 0.00 | 0.12 | 1.00 | 6322 | 6079 |
| Baseline Slope: Type (U): Z-scored TEA | 0.01 | 0.01 | -0.02 | 0.03 | 1.00 | 6822 | 6204 |
| Level Change: Type (U): Z-scored TEA | 0.11 | 0.14 | -0.17 | 0.39 | 1.00 | 8832 | 6215 |
| Slope Change: Type (U): Z-scored TEA | -0.01 | 0.04 | -0.08 | 0.06 | 1.00 | 8781 | 6659 |

**Supplemental Table 8.** Estimated naming accuracy of trained and untrained items for individuals with above average (top) and below average (bottom) sustained attention scores.

|  | Baseline 1 | Baseline 2 | Baseline 3 | **Level Change** | Tx Slope 1 | Tx Slope 2 | Tx Slope 3 | Tx Slope 4 |
| --- | --- | --- | --- | --- | --- | --- | --- | --- |
| Above Average (+1SD) Sustained Attention Score |  |  |  |  |  |  |  |  |
| Trained Items | 15.6% | 16.1% | 16.5% | 60.3% | 64.2% | 67.9% | 71.4% | 74.6% |
| Untrained Items | 16.2% | 17.1% | 18.1% | 29.9% | 32.1% | 34.4% | 36.7% | 39.1% |
| Below Average (-1SD) Sustained Attention Score |  |  |  |  |  |  |  |  |
| Trained Items | 15.3% | 15.9% | 16.7% | 57.8% | 59.5% | 61.1% | 62.8% | 64.3% |
| Untrained Items | 12.6% | 13.4% | 14.2% | 18.5% | 18.7% | 18.8% | 19.0% | 19.2% |

## Supplemental description of Bayesian prior selection

The prior distribution implemented in our models used a mean of zero and standard deviation of 2.5 logits for the time series variables (baseline slope, level change, and slope change), given that we expected the parameter was highly likely to fall within ±5 logits of the mean (Cavanaugh et al., 2022). We included a prior distribution on the intercept using a mean of -1 and SD of 2.5 logits. The intercept prior represents our prior knowledge of likely naming accuracy at the onset of the study baseline based on the stimuli selection criteria consistent with low pre-baseline naming performance (<30% accurate) without tight constraint of the estimate of the intercept. Additional justification supporting the selection of these prior distributions can be found in Cavanaugh et al. (2022) and Evans et al. (2021), where identical prior distributions were implemented. An R-Markdown file detailing model construction and output is provided in the Supplemental Materials.

## References

Beeson, P. M., & Robey, R. R. (2006). Evaluating single-subject treatment research: Lessons learned from the aphasia literature. Neuropsychology Review, 16(4), 161–169.

Bottini, R., & Doeller, C. F. (2020). Knowledge Across Reference Frames: Cognitive Maps and Image Spaces. Trends in Cognitive Sciences, 24(8), 606–619. 10.1016/j.tics.2020.05.008

Boyle, M. (2010). Semantic feature analysis treatment for aphasic word retrieval impairments: What’s in a name? Topics in Stroke Rehabilitation, 17(6), 411–422.

Boyle, M., Akers, C. M., Cavanaugh, R., Hula, W. D., Swiderski, A. M., & Elman, R. J. (2023). Changes in discourse informativeness and efficiency following communication-based group treatment for chronic aphasia. Aphasiology, 37(3), 563–597. 10.1080/02687038.2022.2032586

Boyle, M., & Coelho, C. (1995). Application of semantic feature analysis as a treatment for aphasia dysnomia. American Journal of Speech-Language Pathology, 4, 94–98.

Brady, M. C., Kelly, H., Godwin, J., Enderby, P., & Campbell, P. (2016). Speech and language therapy for aphasia following stroke. Cochrane Database of Systematic Reviews. 10.1002/14651858.CD000425.pub4

Breitenstein, C., Grewe, T., Flöel, A., Ziegler, W., Springer, L., Martus, P., Huber, W., Willmes, K., Ringelstein, E. B., Haeusler, K. G., Abel, S., Glindemann, R., Domahs, F., Regenbrecht, F., Schlenck, K.-J., Thomas, M., Obrig, H., De Langen, E., Rocker, R., … Bamborschke, S. (2017). Intensive speech and language therapy in patients with chronic aphasia after stroke: A randomised, open-label, blinded-endpoint, controlled trial in a health-care setting. The Lancet, 389(10078), 1528–1538. 10.1016/S0140-6736(17)30067-3

Bürkner, P.-C. (2017). **brms**: An *R* Package for Bayesian Multilevel Models Using *Stan*. Journal of Statistical Software, 80(1). 10.18637/jss.v080.i01

Bürkner, P.-C. (2018). Advanced Bayesian Multilevel Modeling with the R Package brms. The R Journal, 10(1), 395. 10.32614/RJ-2018-017

Bürkner, P.-C. (2021). Bayesian Item Response Modeling in *R* with **brms** and *Stan*. Journal of Statistical Software, 100(5). 10.18637/jss.v100.i05

Cavanaugh, R., Quique, Y. M., Dickey, M. W., Hula, W. D., Boss, E., & Evans, W. S. (2022). Practice-Related Predictors of Semantic Feature Verification Treatment for Aphasia. American Journal of Speech-Language Pathology, 31(5S), 2366–2377. 10.1044/2021_AJSLP-21-00296

Cavanaugh, R., Quique, Y. M., Swiderski, A. M., Kallhoff, L., Terhorst, L., Wambaugh, J., Hula, W. D., & Evans, W. S. (2023). Reproducibility in Small- *N* Treatment Research: A Tutorial Using Examples From Aphasiology. Journal of Speech, Language, and Hearing Research, 66(6), 1908–1927. 10.1044/2022_JSLHR-22-00333

Conroy, P., & Scowcroft, J. (2012). Decreasing cues for a dynamic list of noun and verb naming targets: A case-series aphasia therapy study. Neuropsychological Rehabilitation, 22(2), 295–318. 10.1080/09602011.2011.641434

Constantinescu, A. O., O’Reilly, J. X., & Behrens, T. E. J. (2016). Organizing conceptual knowledge in humans with a gridlike code. Science, 352(6292), 1464–1468. 10.1126/science.aaf0941

Creet, E., Morris, J., Howard, D., & Nickels, L. (2019). Name it again! Investigating the effects of repeated naming attempts in aphasia. Aphasiology, 33(10), 1202–1226. 10.1080/02687038.2019.1622352

Diedrichs, V. A., Jewell, C. C., & Harnish, S. M. (2022). A Scoping Review of the Relationship Between Nonlinguistic Cognitive Factors and Aphasia Treatment Response. Topics in Language Disorders, 42(3), 212–235. 10.1097/TLD.0000000000000290

Dignam, J., Copland, D., O’Brien, K., Burfein, P., Khan, A., & Rodriguez, A. D. (2017). Influence of Cognitive Ability on Therapy Outcomes for Anomia in Adults With Chronic Poststroke Aphasia. Journal of Speech, Language, and Hearing Research, 60(2), 406–421. 10.1044/2016_JSLHR-L-15-0384

Dignam, J., Copland, D., Rawlings, A., O’Brien, K., Burfein, P., & Rodriguez, A. D. (2016). The relationship between novel word learning and anomia treatment success in adults with chronic aphasia. Neuropsychologia, 81, 186–197. 10.1016/j.neuropsychologia.2015.12.026

Dignam, J., Rodriguez, A. D., O’Brien, K., Burfein, P., & Copland, D. A. (2023). Early within therapy naming probes as a clinically-feasible predictor of anomia treatment response. Neuropsychological Rehabilitation, 1–24. 10.1080/09602011.2023.2177312

Doogan, C., Dignam, J., Copland, D., & Leff, A. (2018). Aphasia Recovery: When, How and Who to Treat? Current Neurology and Neuroscience Reports, 18(12), 90. 10.1007/s11910-018-0891-x

Edmonds, L. A., Mammino, K., & Ojeda, J. (2014). Effect of Verb Network Strengthening Treatment (VNeST) in Persons With Aphasia: Extension and Replication of Previous Findings. American Journal of Speech-Language Pathology, 23(2). 10.1044/2014_AJSLP-13-0098

Efstratiadou, E. A., Papathanasiou, I., Holland, R., Archonti, A., & Hilari, K. (2018). A systematic review of semantic feature analysis therapy studies for aphasia. Journal of Speech, Language, and Hearing Research, 61(5), 1261–1278.

Epstein, R. A., Patai, E. Z., Julian, J. B., & Spiers, H. J. (2017). The cognitive map in humans: Spatial navigation and beyond. Nature Neuroscience, 20(11), 1504–1513. 10.1038/nn.4656

Evans, W. S., Cavanaugh, R., Gravier, M. L., Autenreith, A. M., Doyle, P. J., Hula, W. D., & Dickey, M. W. (2021). Effects of Semantic Feature Type, Diversity, and Quantity on Semantic Feature Analysis Treatment Outcomes in Aphasia. American Journal of Speech-Language Pathology, 30(1S), 344–358. 10.1044/2020_AJSLP-19-00112

Evans, W. S., Cavanaugh, R., Quique, Y., Boss, E., Starns, J. J., & Hula, W. D. (2021). Playing With BEARS: Balancing Effort, Accuracy, and Response Speed in a Semantic Feature Verification Anomia Treatment Game. Journal of Speech, Language, and Hearing Research, 64(8), 3100–3126. 10.1044/2021_JSLHR-20-00543

Ferguson, A. (1999). Clinical Forum Learning in aphasia therapy: It’s not so much what you do, but how you do it! Aphasiology, 13(2), 125–150. 10.1080/026870399402244

Fillingham, J. K., Sage, K., & Lambon Ralph †, M. A. (2006). The treatment of anomia using errorless learning. Neuropsychological Rehabilitation, 16(2), 129–154. 10.1080/09602010443000254

Foygel, D., & Dell, G. S. (2000). Models of Impaired Lexical Access in Speech Production. Journal of Memory and Language, 43(2), 182–216. 10.1006/jmla.2000.2716

Gilmore, N., Meier, E. L., Johnson, J. P., & Kiran, S. (2019). Nonlinguistic Cognitive Factors Predict Treatment-Induced Recovery in Chronic Poststroke Aphasia. Archives of Physical Medicine and Rehabilitation, 100(7), 1251–1258. 10.1016/j.apmr.2018.12.024

Goldenberg, G., Dettmers, H., Grothe, C., & Spatt, J. (1994). Influence of linguistic and non-linguistic capacities on spontaneous recovery of aphasia and on success of language therapy. Aphasiology, 8(5), 443–456. 10.1080/02687039408248669

Gravier, M. L., Dickey, M. W., Hula, W. D., Evans, W. S., Owens, R. L., Winans-Mitrik, R. L., & Doyle, P. J. (2018). What Matters in Semantic Feature Analysis: Practice-Related Predictors of Treatment Response in Aphasia. American Journal of Speech-Language Pathology, 27(1S), 438–453. 10.1044/2017_AJSLP-16-0196

Helm-Estabrooks, N. (2002). Cognition and aphasia: A discussion and a study. Journal of Communication Disorders, 35(2), 171–186. 10.1016/S0021-9924(02)00063-1

Hope, T. M. H., Seghier, M. L., Leff, A. P., & Price, C. J. (2013). Predicting outcome and recovery after stroke with lesions extracted from MRI images. NeuroImage: Clinical, 2, 424–433. 10.1016/j.nicl.2013.03.005

Hopper, T., & Holland, A. (2005). Aphasia and Learning in Adults: Key Concepts and Clinical Considerations. Topics in Geriatric Rehabilitation, 21(4), 315–322.

Huitema, B. E., & Mckean, J. W. (2000). Design Specification Issues in Time-Series Intervention Models. Educational and Psychological Measurement, 60(1), 38–58. 10.1177/00131640021970358

J. Gabry, R. Češnovar, A. Johnson, & S. Bronder. (2024). cmdstanr: R Interface to “CmdStan” (Version R package version 0.8.1) [Computer software]. https://discourse.mc-stan.org

Kendall, D. L., Oelke, M., Brookshire, C. E., & Nadeau, S. E. (2015). The Influence of Phonomotor Treatment on Word Retrieval Abilities in 26 Individuals With Chronic Aphasia: An Open Trial. Journal of Speech, Language, and Hearing Research, 58(3), 798–812. 10.1044/2015_JSLHR-L-14-0131

Kiran, S., & Bassetto, G. (2008). Evaluating the Effectiveness of Semantic-Based Treatment for Naming Deficits in Aphasia: What Works? Seminars in Speech and Language, 29(1), 71–82. PMC. 10.1055/s-2008-1061626

Kiran, S., & Thompson, C. K. (2019). Neuroplasticity of Language Networks in Aphasia: Advances, Updates, and Future Challenges. Frontiers in Neurology, 10, 295. 10.3389/fneur.2019.00295

Kristinsson, S., Basilakos, A., Elm, J., Spell, L. A., Bonilha, L., Rorden, C., Den Ouden, D. B., Cassarly, C., Sen, S., Hillis, A., Hickok, G., & Fridriksson, J. (2021). Individualized response to semantic versus phonological aphasia therapies in stroke. Brain Communications, 3(3), fcab174. 10.1093/braincomms/fcab174

Laine, M., & Martin, N. (2006). Anomia: Theoretical and clinical aspects. Psychology Press.

Lambon Ralph, M. A., Snell, C., Fillingham, J. K., Conroy, P., & Sage, K. (2010). Predicting the outcome of anomia therapy for people with aphasia post CVA: Both language and cognitive status are key predictors. Neuropsychological Rehabilitation, 20(2), 289–305. 10.1080/09602010903237875

Lazar, R. M., Minzer, B., Antoniello, D., Festa, J. R., Krakauer, J. W., & Marshall, R. S. (2010). Improvement in Aphasia Scores After Stroke Is Well Predicted by Initial Severity. Stroke, 41(7), 1485–1488. 10.1161/STROKEAHA.109.577338

Majerus, S. (2013). Language repetition and short-term memory: An integrative framework. Frontiers in Human Neuroscience, 7. 10.3389/fnhum.2013.00357

Martin, N., & Saffran, E. M. (1999). Effects of Word Processing and Short-term Memory Deficits on Verbal Learning: Evidence from Aphasia. International Journal of Psychology, 34(5–6), 339–346. 10.1080/002075999399666

Massaro, M., & Tompkins, C. (1992). Feature analysis for treatment of communication disorders in traumatically brain-injured patients: An efficacy study. Clinical Aphasiology, 22, 245– 256.

Meyers, J. E., & Meyers, K. R. (1995). Rey Complex Figure Test and Recognition Trial. Psychological Assessment Resources, Inc.

Mirman, D., & Britt, A. E. (2014). What we talk about when we talk about access deficits. Philosophical Transactions of the Royal Society B: Biological Sciences, 369(1634), 20120388. 10.1098/rstb.2012.0388

Nunn, K., Vallila-Rohter, S., & Middleton, E. L. (2023). Errorless, Errorful, and Retrieval Practice for Naming Treatment in Aphasia: A Scoping Review of Learning Mechanisms and Treatment Ingredients. Journal of Speech, Language, and Hearing Research, 66(2), 668–687. 10.1044/2022_JSLHR-22-00251

Peñaloza, C., Martin, N., Laine, M., & Rodríguez-Fornells, A. (2022). Language learning in aphasia: A narrative review and critical analysis of the literature with implications for language therapy. Neuroscience & Biobehavioral Reviews, 141, 104825. 10.1016/j.neubiorev.2022.104825

Plowman, E., Hentz, B., & Ellis, C. (2012). Post-stroke aphasia prognosis: A review of patient-related and stroke-related factors: Aphasia prognosis. Journal of Evaluation in Clinical Practice, 18(3), 689–694. 10.1111/j.1365-2753.2011.01650.x

Pustejovsky, J. E. (2019). Procedural sensitivities of effect sizes for single-case designs with directly observed behavioral outcome measures. Psychological Methods, 24(2), 217–235. 10.1037/met0000179

Quique, Y. M., Cavanaugh, R., Lescht, E., & Evans, W. S. (2022). Applying adaptive distributed practice to self-managed computer-based anomia treatment: A single-case experimental design. Journal of Communication Disorders, 99, 106249. 10.1016/j.jcomdis.2022.106249

Quique, Y. M., Evans, W. S., & Dickey, M. W. (2019). Acquisition and Generalization Responses in Aphasia Naming Treatment: A Meta-Analysis of Semantic Feature Analysis Outcomes. American Journal of Speech-Language Pathology, 28(1S), 230–246. 10.1044/2018_AJSLP-17-0155

Raymer, A. M., Beeson, P., Holland, A., Kendall, D., Maher, L. M., Martin, N., Murray, L., Rose, M., Thompson, C. K., Turkstra, L., Altmann, L., Boyle, M., Conway, T., Hula, W., Kearns, K., Rapp, B., Simmons-Mackie, N., & Gonzalez Rothi, L. J. (2008). Translational Research in Aphasia: From Neuroscience to Neurorehabilitation. Journal of Speech, Language, and Hearing Research, 51(1). 10.1044/1092-4388(2008/020)

Raymer, A. M., & Roitsch, J. (2023a). Effectiveness of Constraint-Induced Language Therapy for Aphasia: Evidence From Systematic Reviews and Meta-Analyses. American Journal of Speech-Language Pathology, 32(5S), 2393–2401. 10.1044/2022_AJSLP-22-00248

Raymer, A. M., & Roitsch, J. (2023b). Word Retrieval Treatments in Aphasia: A Survey of Professional Practice. Aphasiology, 37(7), 954–979. 10.1080/02687038.2022.2063791

Rider, J. D., Wright, H. H., Marshall, R. C., & Page, J. L. (2008). Using semantic feature analysis to improve contextual discourse in adults with aphasia. American Journal of Speech-Language Pathology, 17(2), 161–172.

Robertson, I. H., Ward, T., Ridgeway, V., & Nimmo-Smith, I. (1994). The Test of Everyday Attention (TEA). Thames Valley Test Company.

Robey, R. R. (1998). A Meta-Analysis of Clinical Outcomes in the Treatment of Aphasia. Journal of Speech, Language, and Hearing Research, 41(1), 172–187. 10.1044/jslhr.4101.172

Robinaugh, G., Henry, M. L., Cavanaugh, R., & Grasso, S. M. (2024). Computer-Based Naming Treatment for Semantic Variant Primary Progressive Aphasia With History of Traumatic Brain Injury: A Single-Case Experimental Design. Journal of Speech, Language, and Hearing Research, 67(2), 524–544. 10.1044/2023_JSLHR-23-00289

Rodríguez-Fornells, A., Cunillera, T., Mestres-Missé, A., & de Diego-Balaguer, R. (2009). Neurophysiological mechanisms involved in language learning in adults. Philosophical Transactions of the Royal Society B: Biological Sciences, 364(1536), 3711–3735. 10.1098/rstb.2009.0130

Rose, M. L., Attard, M. C., Mok, Z., Lanyon, L. E., & Foster, A. M. (2013). Multi-modality aphasia therapy is as efficacious as a constraint-induced aphasia therapy for chronic aphasia: A phase 1 study. Aphasiology, 27(8), 938–971. 10.1080/02687038.2013.810329

Sandberg, C. W., & Gray, T. (2020). Abstract Semantic Associative Network Training: A Replication and Update of an Abstract Word Retrieval Therapy Program. American Journal of Speech-Language Pathology, 29(3), 1574–1595. 10.1044/2020_AJSLP-19-00066

Sandberg, C. W., Khorassani, H., Gray, T., & Dickey, M. W. (2023). Novel Participant-Level Meta-Analytic Evidence for AbSANT Efficacy. Frontiers in Rehabilitation Science, 4. 10.3389/fresc.2023.1017389

Seniów, J., Litwin, M., & Leśniak, M. (2009). The relationship between non-linguistic cognitive deficits and language recovery in patients with aphasia. Journal of the Neurological Sciences, 283(1–2), 91–94. 10.1016/j.jns.2009.02.315

Simic, T., Chambers, C., Bitan, T., Stewart, S., Goldberg, D., Laird, L., Leonard, C., & Rochon, E. (2020). Mechanisms underlying anomia treatment outcomes. Journal of Communication Disorders, 88, 106048. 10.1016/j.jcomdis.2020.106048

Simic, T., Rochon, E., Greco, E., & Martino, R. (2019). Baseline executive control ability and its relationship to language therapy improvements in post-stroke aphasia: A systematic review. Neuropsychological Rehabilitation, 29(3), 395–439. 10.1080/09602011.2017.1307768

Stockbridge, M. D., Elm, J., Breining, B. L., Tippett, D. C., Sebastian, R., Cassarly, C., Teklehaimanot, A., Spell, L. A., Sheppard, S. M., Vitti, E., Ruch, K., Goldberg, E. B., Kelly, C., Keator, L. M., Fridriksson, J., & Hillis, A. E. (2023). Transcranial Direct-Current Stimulation in Subacute Aphasia: A Randomized Controlled Trial. Stroke, 54(4), 912–920. 10.1161/STROKEAHA.122.041557

Swiderski, A. M., Quique, Y. M., Dickey, M. W., & Hula, W. D. (2021). Treatment of Underlying Forms: A Bayesian Meta-Analysis of the Effects of Treatment and Person-Related Variables on Treatment Response. Journal of Speech, Language, and Hearing Research, 64(11), 4308–4328. 10.1044/2021_JSLHR-21-00131

Swinburn, K., Porter, G., & Howard, D. (2012). Comprehensive Aphasia Test [Dataset]. American Psychological Association. 10.1037/t13733-000

Thompson, C., & Shapiro, L. (2005). Treating agrammatic aphasia within a linguistic framework: Treatment of Underlying Forms. Aphasiology, 19(10–11), 1021–1036.

Tierney-Hendricks, C., Schliep, M. E., & Vallila-Rohter, S. (2022). Using an Implementation Framework to Survey Outcome Measurement and Treatment Practices in Aphasia. American Journal of Speech-Language Pathology, 31(3), 1133–1162. 10.1044/2021_AJSLP-21-00101

Tuomiranta, L. M., Càmara, E., Froudist Walsh, S., Ripollés, P., Saunavaara, J. P., Parkkola, R., Martin, N., Rodríguez-Fornells, A., & Laine, M. (2014). Hidden word learning capacity through orthography in aphasia. Cortex, 50, 174–191. 10.1016/j.cortex.2013.10.003

Ullman, M. T. (2004). Contributions of memory circuits to language: The declarative/procedural model. Cognition, 92(1–2), 231–270. 10.1016/j.cognition.2003.10.008

Vallila-Rohter, S. (2017). Considering Learning Ability in Language Rehabilitation Plans. Perspectives of the ASHA Special Interest Groups, 2(2), 23–30. 10.1044/persp2.SIG2.23

Vallila-Rohter, S., & Kiran, S. (2013). Non-linguistic learning and aphasia: Evidence from a paired associate and feedback-based task. Neuropsychologia, 51(1), 79–90. 10.1016/j.neuropsychologia.2012.10.024

Varkanitsa, M., Godecke, E., & Kiran, S. (2023). How Much Attention Do We Pay to Attention Deficits in Poststroke Aphasia? Stroke, 54(1), 55–66. 10.1161/STROKEAHA.122.037936

Viganò, S., Rubino, V., Soccio, A. D., Buiatti, M., & Piazza, M. (2021). Grid-like and distance codes for representing word meaning in the human brain. NeuroImage, 232, 117876. 10.1016/j.neuroimage.2021.117876

Votruba, K. L., Rapport, L. J., Whitman, R. D., Johnson, A., & Langenecker, S. (2013). Personality Differences among Patients with Chronic Aphasia Predict Improvement in Speech-Language Therapy. Topics in Stroke Rehabilitation, 20(5), 421–431. 10.1310/tsr2005-421

Wambaugh, J. L., Mauszycki, S., Cameron, R., Wright, S., & Nessler, C. (2013). Semantic feature analysis: Incorporating typicality treatment and mediating strategy training to promote generalization. American Journal of Speech-Language Pathology, 22(2), S334– S369.

Warrington, E. K. (1996). The Camden Memory Tests. Psychology Press.

Watila, M. M., & Balarabe, S. A. (2015). Factors predicting post-stroke aphasia recovery. Journal of the Neurological Sciences, 352(1–2), 12–18. 10.1016/j.jns.2015.03.020

Winans-Mitrik, R., Chen, S., Owens, R., Hula, W. D., Eicchorn, K., Ebrahimi, M., & Doyle, P. J. (2013). Tele-rehabilitation solutions for patients with aphasia [Poster]. Annual Meeting of the Association of Veterans Affairs Speech-Language Pathologists, San Francisco, CA, United States.

Yeung, O., & Law, S.-P. (2010). Executive functions and aphasia treatment outcomes: Data from an ortho-phonological cueing therapy for anomia in Chinese. International Journal of Speech-Language Pathology, 12(6), 529–544. 10.3109/17549507.2011.516840

Yu, C., & Smith, L. B. (2007). Rapid Word Learning Under Uncertainty via Cross-Situational Statistics. Psychological Science, 18(5), 414–420. 10.1111/j.1467-9280.2007.01915.x

